# Using Mobile Phone-based Text Message to Recruit Representative Samples: Assessment of a Cross-Sectional Survey about the COVID-19 Vaccine Hesitation

**DOI:** 10.1101/2022.01.15.22269259

**Authors:** C.I. Sartorao Filho, C.I. Sartorao Neto, A.L.V. Sartorao, D.C. Terribile, R. Mello, B.B. Mello, M.C. Zoqui, D.O. Duarte, L.E.G. Cachoni, V.C.Q. Bisseto, E.A.C. Ribeiro

## Abstract

**Background:** Limited research has examined mobile phone-based platforms for survey recruitment, especially during the COVID-19 pandemic in Brazil. Our objective was to investigate the feasibility and representativeness of mobile phone-based advertisement during a preliminary study about COVID-19 vaccine hesitation in Brazil. Moreover, we evaluate whether the older population can be reached through mobile phone-based platforms of the survey.

**Methods:** We conducted a study in December 2021 based on a preliminary survey about the COVID-19 vaccine hesitation in Assis, Brazil, Sao Paulo state. From a list of the adult population hesitant for the second dose of the COVID-19 vaccine, we sent a mobile phone-based advertisement inviting the participants to answer the survey for one week. The respondent’s data were collected in a Google form platform. The comparison between the target population and the respondents was made using the Chi-squared test and the Welch’s test, using a P-value of .05 as significative.

**Results:** The response rate was 9.99% after one week. The mean age of the respondent group was 33.97 (SD 14.99) and 35.05 (SD 14.19) of the population, with a *P-value* of .192 and a Cohen’s d coefficient of 0.0754, corresponding to a small effect size between groups. We demonstrate that the mobile phone-based survey is a feasible and representative strategy during the pandemics in Brazil. Moreover, the older population respondent was representative.

**Conclusion:** We achieved a representative sample of respondents using the mobile phone-based survey in Brazil. Furthermore, it was representative in all sociodemographic and health characteristics assessed. Finally, these findings suggest the method is a highly feasible and economical means of recruiting for survey research.

## Introduction

Population-based survey research is useful for recruitment and aims for collected samples to be representative of the target population. Otherwise, traditional survey research can be limited by the higher costs, low response rates, and demands more personnel and time be made. (1) In addition, landline telephones are recognized in disuse, limiting their ability for recruitment. Limited research has examined mobile phone-based platforms for survey recruitment, especially in Brazil. (2) Besides, it remains unclear whether the method can recruit a representative sample of older adults (3,4). Therefore, we aimed to investigate whether mobile phone-based text messages can be a feasible, timely, economical, and with minimal human resource commitment means of recruiting a representative sample of adults for a health research survey based on the hesitance for the COVID-19 vaccine in Brazil.

## Methods

At the Educational Foundation of the Municipality of Assis, we conducted a study based on a preliminary survey about the reasons for hesitating or refusing the COVID-19 vaccine in Assis, Brazil, São Paulo state. The Institution Ethical Approval was obtained; CAAE: 51936621.8.0000.8547/Statement: 5.135.036. Consent was obtained before individuals could begin the survey, and if it was not provided, participants were redirected away from the survey. The survey itself was anonymous, and all efforts were made to maintain the confidentiality of individuals who participated. The online text message was used only to advertise and direct interested individuals to the survey link due to COVID-19 vaccine hesitation, hosted with Google Forms. Data were securely kept on password-protected computers and cloud storage accounts. To ensure transparency in the study design and recruitment process, we followed the guidelines for reporting results of internet surveys, describing the checklist details throughout our methods. (5) No incentives were offered for participation. The central theme of our survey was the second dose of COVID-19 vaccine hesitation, focusing on health behaviors, the reasons for, and the self-reported attitude to complete or not the proposed vaccine schedule. A mobile phone-based invitation was sent to all adults from a list provided by the official health authorities, who were delayed more than 30 days for the second dose of the vaccine. Google Forms was used to host the open survey and allow for automatic capture of responses into a spreadsheet. We did not use item randomization or adaptive questioning. There were ten items and four screens, one for the invitation, one for the introduction and consent, one for the survey, and one to thank participants and offer the opportunity to contact us or provide feedback. No completeness check was used, but participants could return to previous pages and change responses. The questions were about the sociodemographic characteristics, the reasons for vaccine hesitation, and the different attitudes toward completing the vaccine proposed scheme.

The official municipality health department provided a list of participants with at least 30 days of delay to the second dose of the COVID-19 vaccine in Assis. The period of observation was from January 01 to November 03, 2021. The health government institution previously agreed to participate in the study. Therefore, we used the spreadsheet containing the name, birth date, gender, address, mobile phone number, date of the first dose, vaccine brand name, the presumed date of the second dose.

Advertisements ran twice a week for each participant’s mobile phone number, during December 10-17, 2018, until the cutoff date for the recruitment. We eligible all the adult participants listed in Assis’s municipality as having the hesitation or refusal for the second vaccine dose more than one month of delay. Invalid, missing data, or inexistent cell phone numbers were excluded.

Data analyses comprised the following three major aspects: data cleaning and checking, descriptive analysis of online text messages ad metrics and costs to determine cost per recruit, and descriptive analyses to provide an overview of our sample and univariate analysis to examine whether the distributions of the sample’s sociodemographic and selected health characteristics were consistent with the target population. All statistical analyses were conducted using Microsoft Excel Version 16.

Concerning response rates, the number of respondents was determined with Google Forms. Unfortunately, Google Forms does not track the number of surveys started—only the number of surveys submitted—and there is no way to prevent or identify when there are multiple entries from the same individual unless they were precise duplicates.

Feasibility was assessed as the ease of conducting research and time commitment and objectively by the costs per recruit. Recruitment costs included advertising costs and administrative costs during the entire recruitment period.

Representativeness of the mobile text message sample was assessed by comparing sociodemographic characteristics of participants respondents with the underlying population, obtained from the census provided by the Health Municipality. These characteristics were age, gender, self-reported comorbidities, type of the first dose vaccine, and the number of delayed days since the recommended second dose date.

We performed Welch’s procedure to compare the two groups (adult population delayed for the COVID-19 vaccine and group of respondents to the mobile phone-based survey), measurement means, and standard deviation. The alpha level was 0.05, two-tails, and the control event rate was 20%. Using an Excel-based calculator, we calculate the mean difference confidence intervals, effect sizes, and Levene’s test of inequality of variance. (6)

The Chi-squared goodness of fit test was used to test if sample data fits a distribution from the population. For statistical analysis, *P*<.05 was considered to indicate rejection of the null hypothesis at the significance level.05. Likewise, *P*>.05 indicates a failure to reject the null hypothesis at the significance level.05, meaning we can assume the sample distribution is consistent or representative with the census distribution.

The magnitude of difference between the respondents’ distribution and the population, for effect size, was calculated and interpreted as per Cohen (7), adopting values of 0.20, 0.50, and 0.80 correspond to small, medium, and large effect sizes. We considered a character to represent the population if the Cohen’s d <0.20 (small effect size).

## Results

There were 4667 vaccine-hesitant adults on the list given by the health authorities from the municipality, delayed for more than 30 days, a mean of 91.39 days (+/- 37.68). In addition, 1120 were excluded due to missing, invalid, or inconsistent mobile phone numbers. Thus, we sent 3547 mobile phone-based text messages. Finally, after one week, we obtained 355 respondents, a rate response of 9.99% after one week of surveillance. Table 1 describes the respondent’s characteristics.

**Table 1.** Respondents Characteristics (N=355)

| Characteristic | Data* |
| --- | --- |
| **Age, Years** |  |
| Mean (SD) | 33.98 (14.99) |
| Range | 18-88 |
| **Age Group, years** |  |
| 18-29 | 179 (50.4) |
| 30-44 | 96 (27.0) |
| 45-59 | 53 (14.9) |
| ≥60 | 27 (7.6) |
| **Gender** |  |
| Female | 153 (43.1) |
| Male | 202 (56.9) |
| **Educational attainment** |  |
| College graduate or above | 80 (22.5) |
| **Have you had COVID-19?** |  |
| Yes (confirmed or clinically suspected) | 37 (10.4) |
| **Self-rated overall health** |  |
| Fair/poor | 56 (15.8) |
| **Brand name of the first COVID-vaccine taken** |  |
| Coronavac | 81 (22.8) |
| Pfizer | 176 (49.6) |
| Astrazeneca | 98 (27.6) |
| **What is your best guess as to whether you will take the second dose of the Coronavirus vaccine?** |  |
| Definitively or probably will take | 286 (80.6) |
| Not sure | 44 (12.4) |
| Probably or definitively will not take | 25 (7.0) |

\*Unless otherwise indicated, data are the respondents' number and percentage (). Percentages may not total to 100 owing to rounding.

Economic considerations suggest that mobile phone text messages are feasible to recruit survey participants. The research was very easy to conduct, manageable by one graduate student as a research assistant committing 14 hours over seven days. This method is also feasible for rapid recruitment of a large sample, from a phone list of 4667 persons, with a mean age of 35.05 years (+/-14.19). We had 355 eligible respondents with a mean age of 33.97 years (+/- 14.99). We had some intangible but inexpensive costs with mobile phone devices and web-based services, and the acquisition of 4 prepaid Subscriber Identification Module cards, for about U$ 10 dollars plus the costs for a private bulk text message service, for U$ 100 dollars. The total amount was around US 110 American dollars or U$ 0.31 per respondent. Thus, the method has a considerably lower cost per respondent.

The population and respondents’ comparison are reported in Table 2, using the Chi-squared tests. There was a statistically significant difference (*P-value:*.001) between the sample and population distributions for the AstraZeneca and Pfizer groups. All other groups were consistent with the population. Likewise, suggests that they are representative of the population.

**Table 2.**
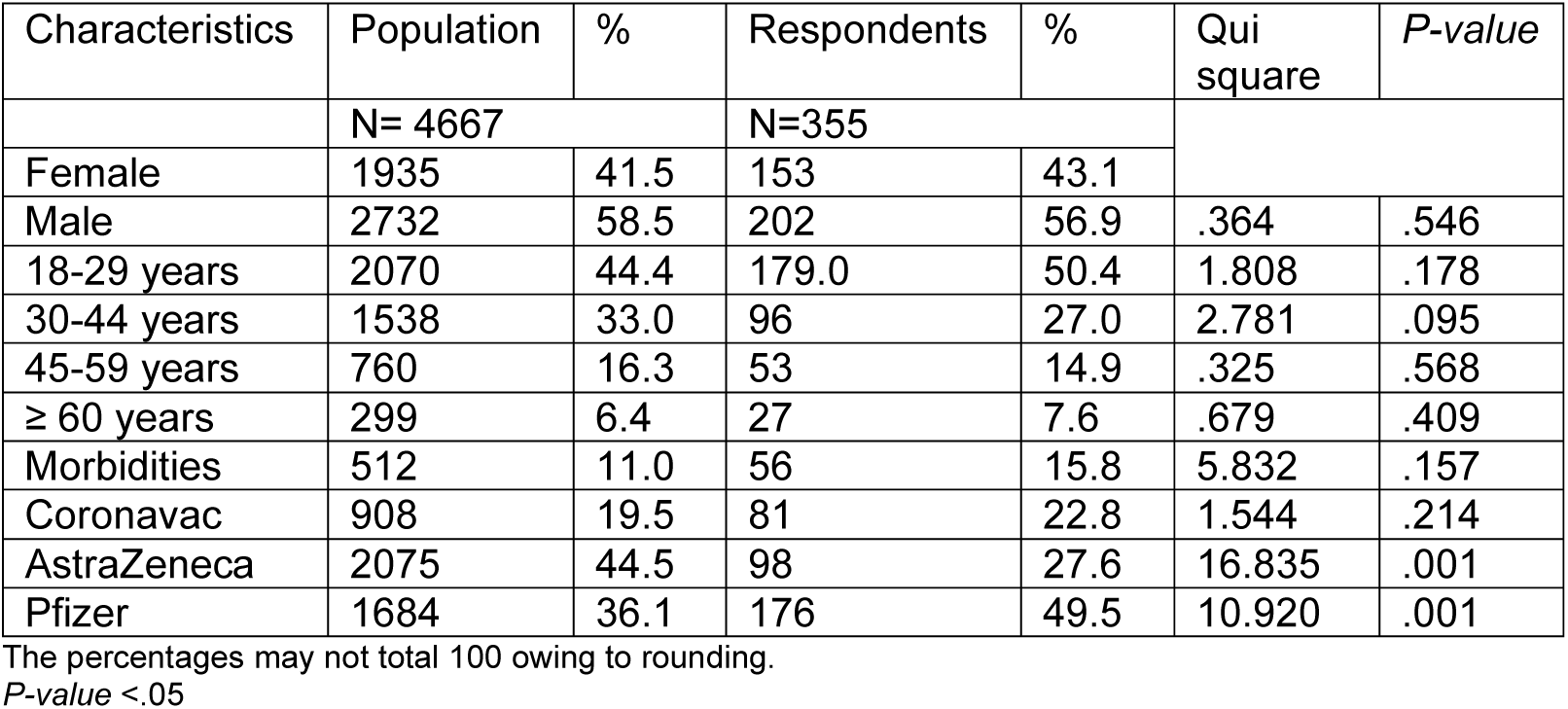
The population and respondents group comparison according to Chi-squared goodness of fit calculation.

Table 3 demonstrated Welch’s test for comparing population and respondent age groups. The Mean age of the respondent group was 33.97 (SD 14.99), and 35.05 (SD 14.19) of the population, with a *P-value* of .192, and a Cohen’s d of 0.0754, corresponding to a small effect size between groups.

**Table 3.**
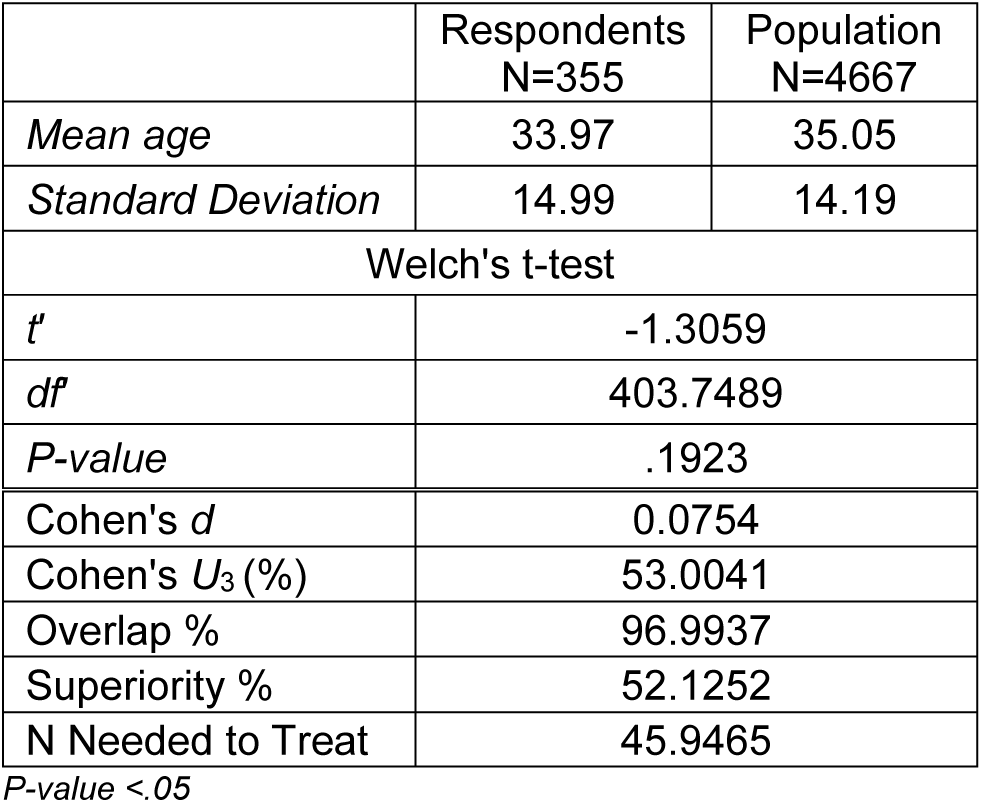
Welch’s test for the comparison between population and respondent age groups.

|  | Respondents<br>N=355 | Population<br>N=4667 |
| --- | --- | --- |
| *Mean age* | 33.97 | 35.05 |
| *Standard Deviation* | 14.99 | 14.19 |
| Welch's t-test |  |  |
| *t* | -1.3059 |  |
| *df* | 403.7489 |  |
| *P-value* | .1923 |  |
| Cohen's *d* | 0.0754 |  |
| Cohen's *U*<sub>3</sub> (%) | 53.0041 |  |
| Overlap % | 96.9937 |  |
| Superiority % | 52.1252 |  |
| N Needed to Treat | 45.9465 |  |

*P-value* <.05

## Discussion

This study investigates whether mobile phone-based advertising can be used to feasibly recruit a representative sample of adults and older adults in a municipality of Brazil to complete a health survey about hesitation for the COVID-19 vaccine. Representativeness was assessed and confirmed by comparing our sample’s numerous sociodemographic and health characteristics with the underlying population. Feasibility was confirmed based on an assessment of costs, ease of use, and recruitment time. Moreover, this is an unprecedented study to investigate mobile phone-based text messages for the recruitment of older adults. The vast majority of the national and international literature on social media recruitment focuses on the youth and young adults, some focusing on middle-aged adult populations. (8–11) The reported good participation of the older respondents’ results should be of considerable interest to academic researchers, community organizations, and governments because they suggest this method can dramatically reduce research barriers with minimal sacrifices to representativeness.

Although the low response rate, the mobile phone-based recruitment sample was representative of the population for age, gender, and self-reporting comorbidities. Targeting by age and gender was effective in increasing representation. We must assume an over-representation of people with higher levels of education, but targeting advertisements proved both costly and ineffective at improving the representation of participants with lower levels of educational attainment. The response rates of all survey methods decreased over the years, possibly due to a proliferation of web-based questionnaires. (12)

Our reporting contributes to the evidence-based use of online text message surveys for health research. This recruitment method works effectively in the context of the Assis city population. Furthermore, we have shown that online text messages can recruit older adults to research surveys, where the vast majority of previous research on this subject has only considered younger populations. (13)

Caution should be used to consider the first dose vaccine groups from AstraZeneca and Pfizer results. Although significant, there is a potential possibility that these groups have a particular related interest to respond or not to the survey. Therefore, we emphasize the need to proceed with further investigations to elucidate these findings.

The research was very easy to conduct and manageable by one graduate student committing 14 hours per week as a research assistant. This method is also feasible for the rapid recruitment of a large sample, with 355 respondents being recruited in only seven days. A major advantage over other recruitment methods is that it was easy to use targeted advertisements to improve a sample’s representativeness and recruit hard-to-reach populations. There was also essentially no footwork involved, such as putting up recruitment posters, and we believe there was less selection bias than using email listservs or snowball sampling. (14,15) Otherwise, there were no risks for the online researchers and the target population in transmitting diseases through face-to-face surveys, especially during the pandemics. (16)

We also found that a mobile phone-based survey is an economical means of recruiting participants for survey research. It has the potential to be a relatively less expensive and timely method of collecting survey information. (17) But, again, maybe due to the higher population interest in the vaccine as an issue that affects everyone.

Limitations of our study were based mostly on subjective interpretations about the feasibility. As our analysis compares representativeness with the population, it is impossible to directly compare the representativeness of samples recruited with mobile phone-based with different recruitment methods. Another perceived limitation was the population’s distrust in receiving non-official and unexpected phone messages. Moreover, we can not account for the real number of delivery messages due to limitations as internet failure, missing mobile phone numbers, or interrupted plan services, especially in the low-income population.

We believe there are many potential applications of mobile phone-based advertising for researchers, governments, and community-based organizations who wish to learn about the populations they serve. Besides, we believe that these institutions should previously officially advertise the target population to promote a wider adhesion and confidence to respond to the survey.

## Conclusion

We achieved a representative sample of respondents using the mobile phone-based survey in Brazil. Furthermore, it was representative in all sociodemographic and health characteristics assessed. Finally, these findings suggest the method is a highly feasible and economical means of recruiting for survey research.

## Data Availability

All data produced in the present study are available upon reasonable request to the authors

